# Nuclear myocardial perfusion studies do not predict vascular or myocardial architecture in advanced heart failure

**DOI:** 10.1101/2020.12.18.20244293

**Authors:** Stephen Farris, Creighton Don, Laurie Soine, William Lombardi, James Caldwell, April Stempien-Otero

## Abstract

**Objectives:** To determine predictors of tissue that would benefit from improved blood flow in subjects with advanced heart disease.

**Background:** Advanced regeneration and percutaneous revascularization strategies including revascularization of chronic total occlusions (CTO) are being used to potentially restore myocardial function in the growing population of patients with advanced heart disease. Predictors of myocardial recovery in this population have not been established.

**Methods:** Cardiac tissue was collected from 27 subjects undergoing LVAD or cardiac transplantation who had nuclear perfusion imaging prior to procedure. Histologic analysis for fibrosis, vascular density and inflammation was performed and correlated with nuclear perfusion images from the same areas. In a subset of subjects, correlations of nuclear perfusion with TIMI flow by coronary arteriography was also performed.

**Results:** Areas of both normal and abnormal epicardial coronary blood flow had a large variation in radiotracer uptake. There were no statistically significant correlations between uptake of radiotracer at rest and degree of cardiac fibrosis, vascular density or inflammation and the matched area of myocardium. Fibrosis varied from 5-50% in areas with severe defects and areas of normal radiotracer uptake. Endothelial density correlated with inflammation in end-stage heart disease.

**Conclusions:** Radiotracer uptake poorly predicted both areas of severe fibrosis that will not benefit from increased blood flow and areas of minimal fibrosis that may recover. Correlation of inflammation and endothelial density support a hypothesis that inflammatory cytokines augment vascularity in myocardium. Novel strategies to improve function in advanced heart disease are in need of better markers of recovery.

## Introduction

The prevalence of ischemic cardiomyopathy is dramatically increasing due to both success with early intervention in myocardial infarction and an increase in an aging population with a worsening comorbidity profile (1, 2). Although transplant is the best treatment for advanced heart failure caused by ischemic cardiomyopathy, it is limited resource. Furthermore, patients with ischemic cardiomyopathy are frequently not ideal candidates for transplant as they are older, have frequently had previous sternotomies and have more risk factors for poor outcomes with transplant such as peripheral vascular disease (1, 3, 4). As a consequence, novel therapies specific for ischemic cardiomyopathy have been in development. These include percutaneous strategies to revascularize total occlusions, less invasive surgical strategies, more comfort with higher risk surgery with LVAD back up, and truly novel therapies such as stem cell therapies.

Defining who will benefit from these novel therapies is a challenge. Patients with ischemic cardiomyopathy have limitations in both macro- and micro-vascular blood flow. Furthermore, years of remodeling and strain result in additional cardiomyopathic changes including fibrosis and inflammation. Classically, these novel therapies are directed at improving contractile function by improving oxygen delivery to “hibernating” myocardium. However, recent data suggest that improving microvascular blood flow can have beneficial effects on other facets of myocardial remodeling such as matrix turnover, inflammation and oxidative stress. It is clear that well designed clinical trials are needed to determine if, how, and in whom these novel revascularization therapies are of benefit.

Integral to both designing clinical trials and treating individual patients are methodologies to define the location of recoverable tissue. Cardiac MRI provides the most detailed assessment of fibrosis, however, most patients with ischemic cardiomyopathy have implanted defibrillators that create significant artifact with current techniques. Thus, perfusion imaging strategies designed to look at both blood flow and the presence of viable myocardium have been the gold standard for determining revascularization strategies (5-7). Unfortunately, echo perfusion imaging requires expertise beyond most centers, and the performance of nuclear imaging in this population is uncertain as the original animal validation studies were performed in the setting of experimental myocardial infarction without significant ventricular remodeling(8) (9). Furthermore, we used SPECT in a study of injection of cell therapeutics to areas with moderate ischemia (25-50% uptake) but found a wide range of fibrosis in explanted tissue(10).

In light of these data, we used a novel experimental opportunity to determine if myocardial radiotracer uptake can identify tissue that may benefit from therapies to improve macro or micro vascular disease in advanced ischemic cardiomyopathy. Patients with ischemic and non-ischemic cardiomyopathies who had undergone SPECT or PET imaging as a part of work up for advanced heart failure therapies had tissue collected at the time of these therapies (LVAD implantation or cardiac transplantation) and detailed tissue characteristics were compared to imaging findings. We found no correlation between uptake of nuclear tracer and fibrosis or vascular density. These data suggest that this imaging modality should not be used to include or exclude patients with ischemic cardiomyopathy from novel revascularization strategies.

## METHODS

### Study setting and Population

Subjects gave informed consent and were prospectively enrolled at the University of Washington Medical Center prior to left ventricular device (LVAD) implantation (as either bridge to transplant or destination therapy) or primary transplant/Total Artificial Heart (SynCardia) implantation. All study protocols were approved by the University of Washington IRB and adhered to the Helsinki Principles for Human Subjects Research.

### Clinical Characteristics

Subjects underwent routine clinical care with established protocols at the University of Washington. Demographics, clinical parameters, and nuclear perfusion imaging (as clinically indicated) were obtained prior to LVAD implantation or heart transplantation/ Total Artificial Heart (SynCardia) implantation.

### Myocardial Perfusion Imaging

Myocardial perfusion imaging (10 thallium^201^ SPECT, 12 Technicium^99^ SPECT and 5 PET) were acquired during routine clinical care and analyzed in the nuclear cardiology lab using Cardiac QGS software viewed on a GE Xeleris work station. Resting myocardial perfusion was quantified in segments 7 and 12 (for the anterolateral wall) and 17 (for the apex) per the standard 17-segment model.(11) Radiotracer uptake defects were determined using semi quantitative distribution of radiotracer into quartiles of percent normal (0 = normal, 1 = 0 – 25%, 2 = 25-50%, 3 = 50-75% and 4 = 75-100% perfusion defect). Radiotracer uptake was evaluated in a standard and blinded fashion with respect to subject and etiology of cardiomyopathy by S.F. and L.S. There was agreement for 33/37 regions of interest between L.S. and S.F., consensus was reached for the 4 differing regions. (Figure 1)

**Figure 1.**
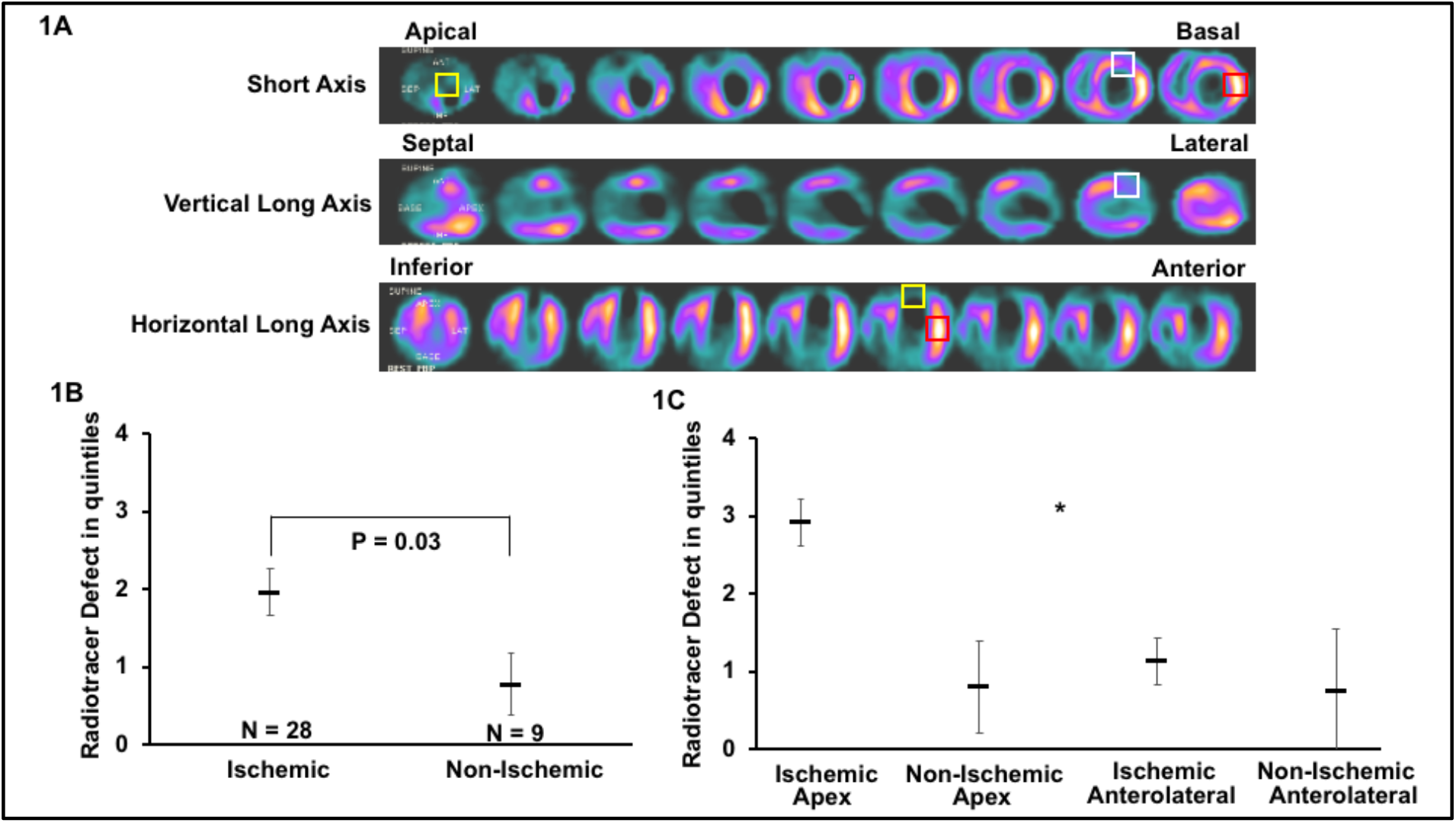
Perfusion Analysis in. **(A)** Example of resting perfusion study on a subject with ischemic cardiomyopathy using Tc-99 MIBI. SPECT images were obtained preoperatively during routine clinical care. Radiotracer uptake in the basal anterolateral, apical and normal reference segments (white, yellow and red boxes, respectively) were determined using orthogonal axes. Defects were divided into quintiles in the usual semi-quantitative fashion (0 = normal, 1 = 0 – 25%, 2 = 25-50%, 3 = 50-75% and 4 = 75-100% perfusion defect) and displayed as the group mean ± SEM. Only regions with acquired tissue and histologic comparison are presented and stratified as ischemic vs. non-ischemic **(B)** or combined with region of interest **(C). \*** = P < 0.01 for the interaction between cardiomyopathy etiology and region of interest by ANOVA on ranks.

### Tissue Processing and Histologic Studies

Full thickness pieces (1-2g) of left ventricular anterolateral wall or apical core myocardial tissue were obtained in the operating theater and underwent immediate formalin fixation followed by paraffin embedment for histology. Myocardial tissue was characterized for capillary density, macrophage density and cardiac fibrosis as previously described.(12) Quantification was performed in a blinded fashion with respect to subject and etiology of cardiomyopathy using automated image analysis by the University of Washington Histology Core or manual analysis using Image-Pro Analyzer 7.0 (Media Cybernetics, Inc.) in our lab.

### Statistical Analysis

Demographic and clinical data are presented as median or mean as indicated ± SEM. Spearman Rank Order Correlation, Student T-tests, and ANOVA tests are used when appropriate. Unless stated otherwise, data is normally distributed.

## RESULTS

### Subject Characteristics

Thirty-seven myocardial tissue samples with corresponding preoperative nuclear imaging were obtained from twenty-seven subjects. Subject median age was 60 years with most being male. Tissue was obtained primarily from subjects with ischemic cardiomyopathy (74%). Full thickness myocardial samples were acquired in a consistent fashion from the apical core at the time of LVAD insertion (24), apex and anterolateral wall at the time of primary transplant or TAH implantation (3), and anterolateral wall at transplantation after mechanical support (10). We have previously shown in a larger cohort that there are no changes in the measured histologic parameters with LVAD unloading. There were no differences in subject age, gender, medication use or length of mechanical support with respect to etiology of heart failure (Table 1). (10)

**Table 1.** Subject Characteristics.

|  | N (%) |
| --- | --- |
| Total Enrolled | 27 |
| Age (median) | 60, range 33-72 |
| Male | 23 (85%) |
| Ischemic | 20 (74%) |
| Mechanical Support | 24 (89%) |
| Ejection Fraction | 22.7 ± 11.1 % |

### Characteristics of myocardial perfusion imaging in advanced heart disease

For the myocardial perfusion images, apical and anterolateral wall segments corresponding to tissue collection sites were easily identified (Figure 1A). Mean radiotracer uptake defects (in quintiles and as compared to normal region) were worse in ischemic vs. non-ischemic myocardium as expected (2 ± 0.3 vs 0.8 ± 0.4, respectively, P = .04, Figure 1B). Consistent with a history of anterior myocardial infarction being associated with development of advanced heart failure, apical segments had significantly less radiotracer uptake vs. anterolateral wall (2.3 ± 0.4 vs. 1.1 ± 0.3, respectively, P < .01) (Figure 1C).

Most segments analyzed demonstrated normal epicardial coronary flow on paired angiograms (30 out of 35, Supplemental Figure 1A). Evidence of reduced flow was seen in five segments with severe tracer defects but vessels with normal flow corresponded to regions with severe tracer defects as well (Supplemental Figure 1B, C).

### Capillary density is not associated with radiotracer uptake at rest

Endothelial capillary density was determined by PECAM-1 staining and compared to resting myocardial radiotracer uptake. We found no significant correlation between capillary density and radiotracer uptake for all apical and anterolateral regions (2A) (n = 5 - 12 samples per quintile of defect, P = 0.5 by ANOVA). Median capillary density was nearly identical between areas without perfusion defects as those with severe perfusion defects (0.67 vs 0.4 % PECAM positive area, Figures 2B, C). Indeed, the two samples with the highest capillary densities correlated with significant perfusion defects and tissue samples with very little capillary density were spread across the perfusion defect quintiles (Figure 2D). As myocardial perfusion imaging may be less applicable in non-ischemic cardiomyopathy, we examined but failed to find an association between capillary density and radiotracer uptake in ischemic tissue (Supplemental Figure 2A). There were no statistical correlations between location of samples (apical vs anterolateral) or etiology of heart failure. There were no significant interactions between perfusion imaging technique and capillary density.

**Figure 2.**
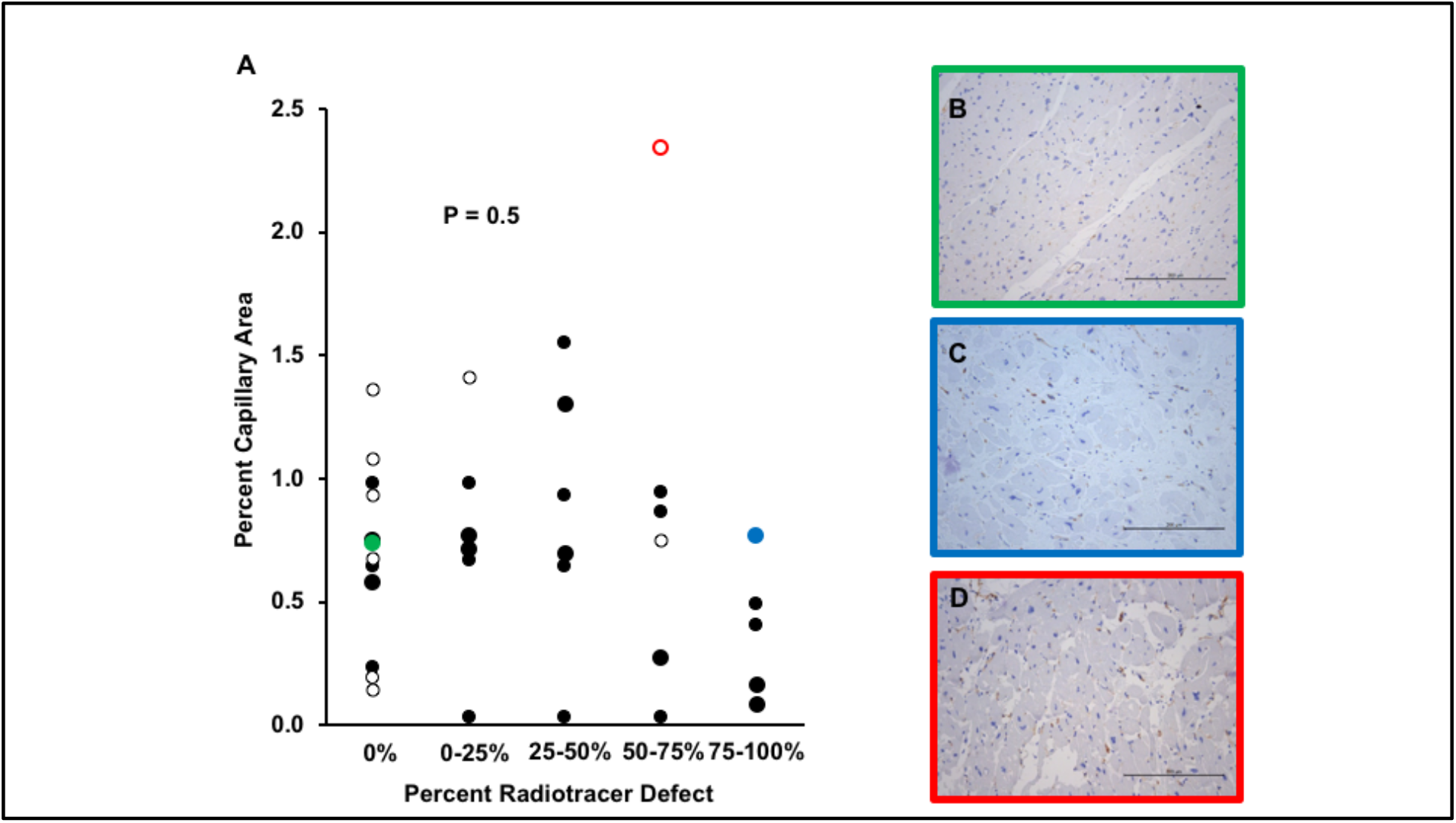
Resting myocardial perfusion does not correlate with myocardial capillary density. **A**. Individual data points are represented by closed circles (ischemic) and open circles (non-ischemic) (P = 0.5 by ANOVA). **B-D**. Representative micrographs at 20x in which box outline color corresponds to color of highlighted data point. Note that regions with identical capillary density can have **B**. low or **C**. high perfusion defects. **D**. represents area with highest capillary density which of note also had significant perfusion defect. Capillary density is quantified as percent positive area stained for mouse anti-human platelet endothelial cell adhesion molecule (PECAM)-1/CD31 with brown area corresponding to endothelial wall. Tissue is counterstained with hematoxylin. Bar = 200 uM.

### Cardiac fibrosis is not associated with radiotracer uptake at rest

Myocardial fibrosis severity was determined by picrosirius red staining and calculated as the percent fibrotic area. There was no correlation between resting myocardial radiotracer uptake in corresponding regions of high or low fibrosis (Figure 3A). As expected, many areas of low fibrosis had normal to nearly normal radiotracer uptake. However, the converse was not necessarily true: fibrosis content varied dramatically from 6.8 to nearly 50% of the tissue in areas read has having no evidence of a perfusion defect. Examples of this discordance were seen in both ischemic (Figure 3 B, C) and non-ischemic (Figure 3 D, E) tissues. There was also no significant association between radiotracer uptake and severity of myocardial fibrosis in the ischemic cohort (Supplemental Figure 2B).

**Figure 3.**
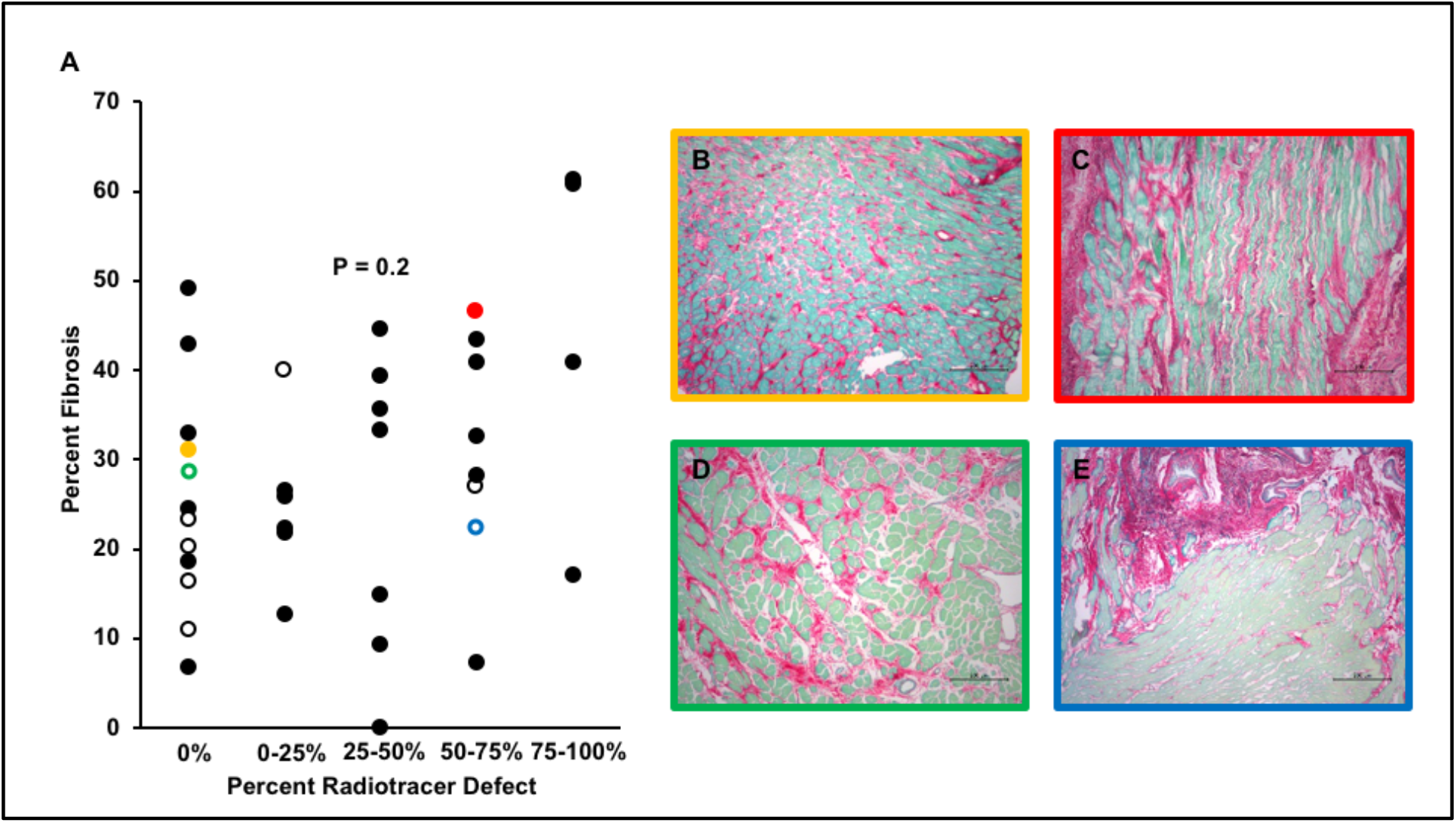
Resting myocardial perfusion does not correlate with myocardial fibrosis. **A**. Individual data points are represented by closed circles (ischemic) and open circles (non-ischemic) (P = 0.2 by ANOVA). **(B-E)**. Representative micrographs at 10x corresponding to regions with varying low perfusion defects. There is minimal difference in fibrosis between areas with no perfusion defect versus severe perfusion defects in subjects with either ischemic **(B, C)** or non-ischemic **(D, E)** heart disease. Fibrosis density is quantified as percent positive area stained for picrosirius red with pink area corresponding to collagen. Tissue is counterstained with fast green. Bar = 200 uM.

### Inflammation is not associated with radiotracer uptake at rest

Because inflammatory cells can take up radiotracer,(13-15) we tested the hypothesis that increased levels of inflammation were associated with increased uptake. Inflammatory cells, primarily macrophages, were identified by immunostaining for CD68. There was no association between the density of inflammatory cells and uptake of radiotracer (Figure 4A). As we have previously shown macrophages were quite prominent in many samples from these subjects (Figure 4B).

**Figure 4.**
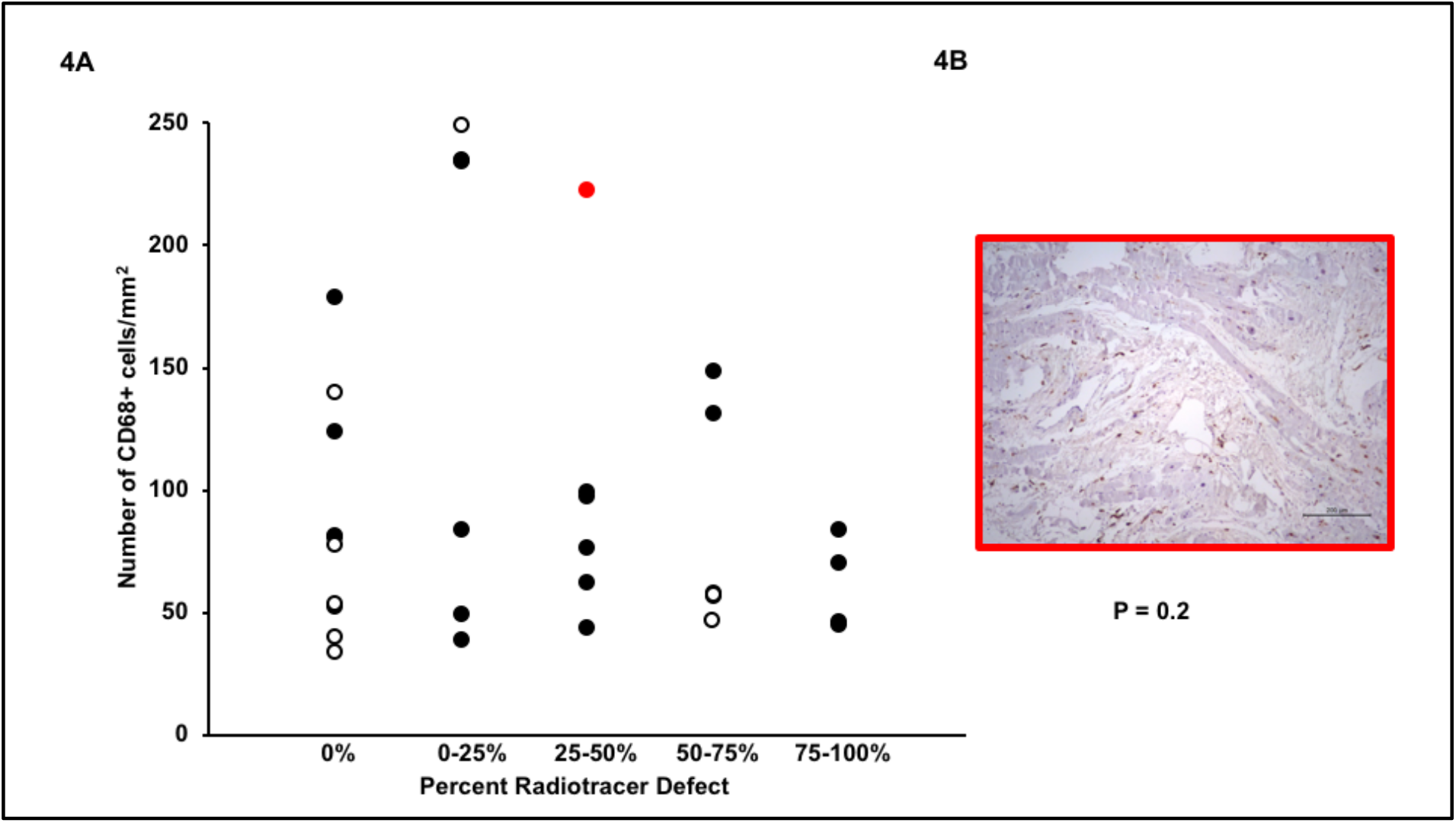
Resting myocardial perfusion does not correlate (P = 0.2 by ANOVA) with myocardial inflammation. Individual data points are represented by closed circles (ischemic) and open circles (non-ischemic) **(A)**. Representative micrograph at 10x of CD68 staining (brown cells) **(B)**. Macrophage density is quantified as # CD68+ cells/mm^2^ tissue. Tissue is counterstained with hematoxylin. Bar = 200 uM.

### Endothelial density is associated with macrophage inflammation in end-stage heart failure

To determine if increased density of capillaries was associated with healthier myocardium, we tested the hypothesis than endocardial density was associated with less inflammation and fibrosis, both hallmarks of heart failure of any etiology. Surprisingly, we found that increasing capillary density correlates highly significantly with increasing inflammation as determined by macrophage density (Figure 5A R= 0.73, P < 0.0001) however there is no association with activated myofibroblasts (Figure 5B R = -0.18, P 0.3) or total fibrosis burden (Figure 5C R = - 0.01, P =0.9). There are no interactions with respect to tissue location or etiology of heart failure with respect to the association of capillary density and inflammation, myofibroblast and fibrosis density (data not shown).

## DISCUSSION

Barriers to myocardial recovery in ischemic cardiomyopathy are many and include a relatively poor understanding of myocardial remodeling in patients with advanced disease. As more patients survive longer past their initial ischemic events, extensive remodeling of scar, ischemic and non-ischemic myocardium occurs. At the same time, recurrent ischemic events and the need for revascularization continue. Radionuclide perfusion imaging is often used to determine the recoverability of myocardium in these patients. Interpretation of these images is based on assumptions of relatively distinct areas of scar, functioning myocardium, and hibernating myocardium. However, in patients with long standing ischemic disease, chronic increases in wall tension and neurohormonal stimulation lead to a heterogenous tissue environment with fibrosis and inflammation in both ischemic and uninjured myocardium (16, 17).

We used a unique circumstance-patients undergoing cardiac transplantation or implantation of durable mechanical support-to directly analyze tissue in this patient population and correlate it with pre-operative perfusion imaging. Our major finding was that this imaging modality is a very poor predictor of myocardial tissue characteristics in an advanced heart failure population. Specifically, increased uptake of tracer was not associated with increased capillary density and decreased uptake not associated with the amount of cardiac fibrosis. In sum, these data demonstrate that even “normal” uptake of radiotracer does not indicate that the underlying tissue represents viable myocardium.

These data are consistent with and may also explain the decades-old observation that patients with non-ischemic cardiomyopathy have perfusion defects on both SPECT and PET imaging(18). Although scans from subjects with ischemic disease had more perfusion defects than those with non-ischemic disease, mostly in the anterior wall, there were no other differences between the groups. Likewise, tissue from subjects with non-ischemic disease had areas of dense fibrosis indistinguishable from ischemic scar. This overlap supports the hypothesis that advanced cardiac remodeling has common pathways between ischemic and non-ischemic cardiomyopathies (19).

Our data differ from those of earlier groups who evaluated the association of myocardial perfusion with histologic analyses. (20) In a cohort of thirty subjects with significant LAD coronary disease undergoing coronary artery bypass grafting, Maes et al found a statistically significant association of preoperative Tc-99 MIBI uptake and percent fibrosis (R 0.78). However, only ten percent of their subjects had heart failure and the average ejection fraction was over forty-five percent.(21) In a smaller cohort of twenty subjects with coronary artery disease and an ejection fraction of ∼ 30%, Shimoni et al tested the hypothesis in pre-CABG subjects that microvascular perfusion by contrast echocardiography would be associated with histologic architecture. They found a moderate association of peak myocardial contrast intensity and capillary area (R = 0.64) and fibrosis (R = -.45).(5) Of note, their cohort had few biopsy specimens with greater than thirty percent fibrosis, consistent with less severe myocardial disease. More consistent with our data, in thirteen subjects with ischemic cardiomyopathy undergoing transplantation, Shirani et al showed a correlation of Thallium uptake to fibrosis (R – 0.7) in regions with transmural infarction that was lost in non-transmural infarcted segments.(19)

Our data do not reveal a single cause for this discrepancy between SPECT uptake and tissue characteristics. However, the most significant association that we found in this study was between inflammatory cells and capillaries. Areas with the highest density of inflammatory macrophages (measured as CD68 positivity) also had the highest density of capillaries. Inflammatory macrophages are known to potentiate angiogenesis through multiple mechanisms including elaboration of pro-angiogenic growth factors (for review see Corliss et al)(22). The increase in capillaries in macrophage-rich segments could result in increased delivery of tracer that could overestimate the amount of viable myocardium in these areas(9). On the other hand, inflammation is also associated with defects in mitochondrial function which could decrease uptake of tracer by viable cardiomyocytes (23). The balance of these factors in a given area of myocardium likely contributes to the unpredictable uptake of tracer and poor predictive value for the study.

This study has several limitations. Although our data indicate that SPECT uptake should not be used to assess myocardial viability in subjects with advanced ischemic cardiomyopathy, very few of the areas that we assessed were supplied by a stenotic or occluded vessel that would be a target for revascularization. Secondly, many argue that PET has improved sensitivity over SPECT for imaging viable myocardium. Only a few of our subjects had PET studies, however, they showed similar poor predictive value. Furthermore, the physiology of PET studies, in particular–measuring cell metabolism with FDG–is as likely to target areas of inflammation as viable myocardium. Finally, although our previous studies demonstrated consistent tissue characteristics throughout the core of tissue collected, areas evaluated histologically could be non-representative of the entire region of interest.

Novel therapeutics are increasingly being used to treat ischemic cardiomyopathy including complex revascularization (both percutaneous and surgical) and cell therapeutics. Studies to determine the effectiveness of these procedures will depend on targeting of these therapies to areas of poorly functioning but viable myocardium. Furthermore, these areas must have similar characteristics from subject to subject. Our data demonstrate that the use of radionuclide perfusion imaging to identify these areas is flawed. If we wish to test the hypothesis that these areas will benefit from a novel therapeutic strategy, then direct measures of fibrotic content such as MRI are likely more precise. Although many of these patients have ICD’s in place that hamper interpretation with current modalities, our data support that research on improved protocols to counter ICD artifacts would be of extremely high yield in this growing population.

## Data Availability

All data is within manuscript

## ACKNOWLEDGEMENTS

We would like to acknowledge the assistance of Susanne Steffes, RN, DNP with patient recruitment and Deri Helterline MS with histology.

---

**Supplementary Figure 1.**
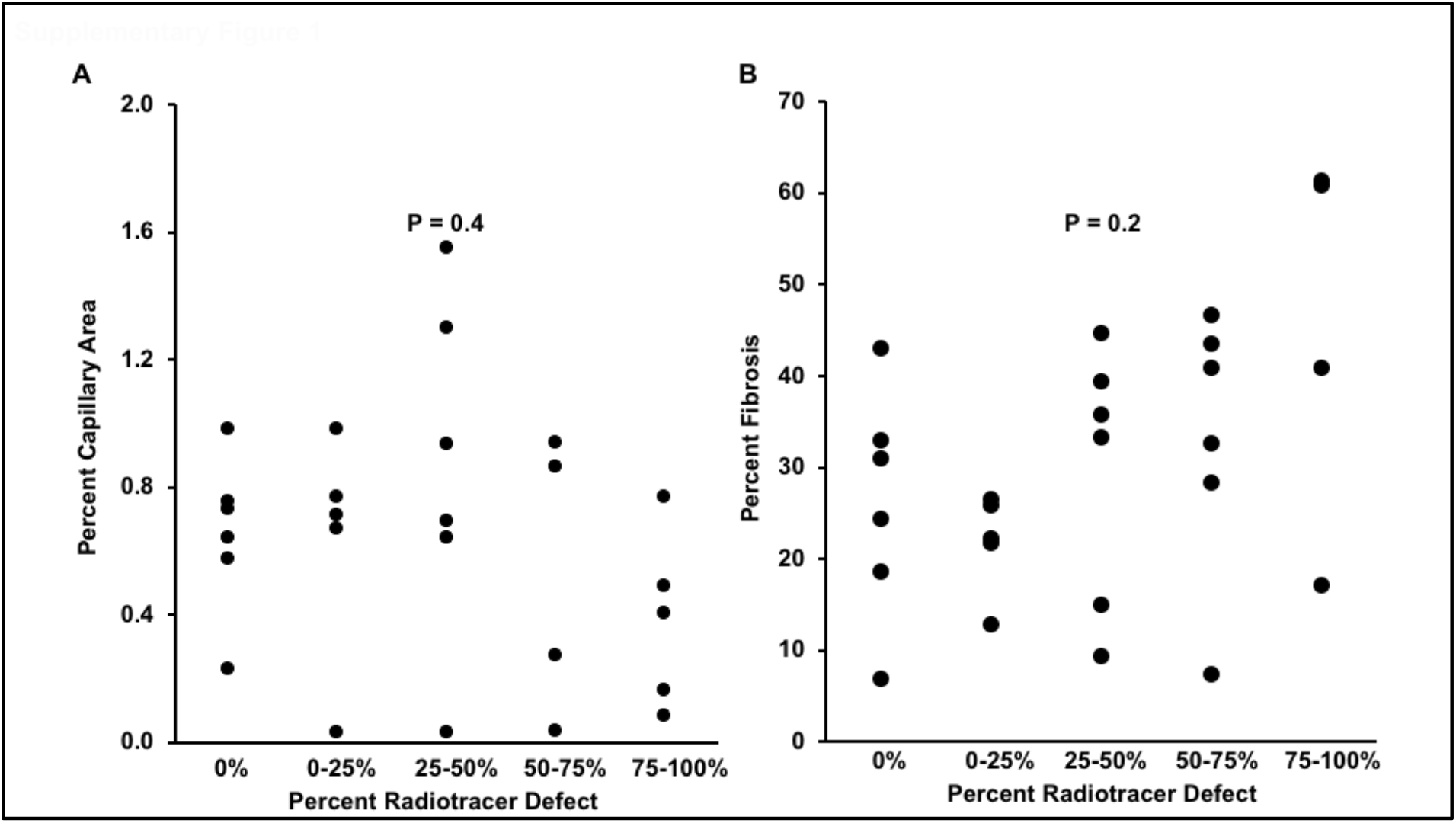
Resting myocardial perfusion does not correlate with capillary density (P = 0.4 by ANOVA) **(A)** or myocardial fibrosis (P = 0.2 by ANOVA) **(B)** in ischemic tissue. Individual data points are represented by circles.

